# Increasing frequency of secondary dengue infections in sequential outbreaks (2016-2024). Clinical impact and diagnostic challenges

**DOI:** 10.64898/2026.05.29.26354405

**Authors:** Sonia L Espindola, Matías Javier Pereson, José Martín Lema, Analía Kachuk, Graciela M Carballo, Natalia Aloisi, María Noel Badano, Marcos Miretti, Federico A Di Lello, Patricia Baré

## Abstract

Successive dengue virus (DENV) outbreaks can progressively reshape population immunity influencing disease expression and diagnostic performance.

**Objectives:** The aim was to evaluate the impact of secondary infections across sequential outbreaks on clinical severity, serotype dynamics and diagnostic concordance.

**Methods:** This retrospective study analyzed 976 febrile-stage samples from three sequential outbreaks in Misiones, Argentina. For serotyping and clinical analyses, 869 viremic samples confirmed by at least one direct method were included (2016: n=512; 2019: n=148; 2024: n=209). Additionally, 318 samples, including 107 non-viremic cases, were used to compare NS1 rapid diagnostic tests (NS1 Ag) and RT-PCR. Viral serotyping and clinical and laboratory markers of disease severity were evaluated.

**Results:** Secondary infections increased from 31.05% (2016) to 43.24% (2019) and 53.87% (2024) (p<0.0010). Serotype distribution shifted from DENV-1 predominance in 2016 (95.12%), DENV-1/DENV-4 co-circulation in 2019 (60.71%/39.29%), and DENV-2 predominance in 2024 (97.60%). Secondary infections were associated with more severe disease manifestations, particularly in 2024, with higher hematocrit (p=0.0120) and hemoglobin (p=0.0080), lower white blood cells (p=0.020) and platelet counts (p=0.0030), and elevated AST (p=0.0007) and ALT (p=0.0130). Concordance between NS1 Ag and RT-PCR was lower in secondary infections (κ=0.457 vs κ=0.759, p=0.0013).

**Conclusions:** The rising frequency of secondary infections may affect both clinical severity and diagnostic performance during outbreaks. The clinical impact was more evident in 2024, likely associated with the introduction of a new serotype. These findings highlight the need for optimized surveillance and diagnostic strategies to improve case detection and patient management during epidemics.

## INTRODUCTION

Dengue fever is a systemic mosquito-borne viral disease caused by dengue virus (DENV), primarily transmitted by *Aedes aegypti*. It has become the most widespread arboviral infection globally, with an estimated 390 million infections annually [1]. In 2024, the Americas reported an unprecedented surge exceeding 14 million cases and more than 12,000 deaths [2]. According to historical records, the province of Misiones in Argentina has experienced three successive dengue outbreaks, since 2016 [3–8]. However, the proportion of secondary DENV infections and their impact upon clinical manifestations and diagnostic performance remain unexplored.

Secondary infections are a key driver of disease burden. The risk is shaped by serotype prevalence, co-circulation patterns, and the host immune profile, including prior exposure and antibody titers that influence protection or enhancement [9–12]. Despite some controversy, secondary infections are recognized as an important determinant of clinical severity [13].

Biochemical parameters can reflect dengue progression. Elevated hematocrit indicates plasma leakage; thrombocytopenia and leukopenia are associated with progression to hemorrhagic shock and increased hepatic enzymes suggest organ involvement and poorer prognosis. Together, these markers help identify warning signs of DENV infection.

During outbreaks, rapid diagnostic tests (RDT), typically immunochromatographic assays detecting NS1 antigen (NS1 Ag) and/or IgM/IgG, are widely used because they require minimal infrastructure and provide quick results [14–16]. During the acute febrile phase, guidelines recommend combining antigen detection, serology, and RT-PCR (when available), interpreted according to illness timing and clinical presentation [16]. However, their performance in secondary dengue infections remains poorly defined, particularly during large-scale outbreaks. This study aimed to determine serotype dynamics and how secondary dengue infections influence clinical severity and affect the reliability of diagnostic tools during large successive outbreaks.

## MATERIALS AND METHODS

### Samples and Study Design

This retrospective study was conducted on 976 febrile-stage samples obtained from 2016 to 2024 during sequential DENV outbreaks in the city of Posadas, Misiones (northeastern Argentina). For the analysis of serotyping and clinical manifestations, we included 869 viremic samples from 2016, 2019, and 2024. These samples were positive by at least one direct diagnostic method (NS1 antigen, RT-PCR, or both). Routine clinical and laboratory data were retrieved from the laboratory system and medical records.

A subgroup of 318 serum samples from the 2016 outbreak, including 211 viremic and 107 non-viremic samples, were evaluated to compare assays’ performance and their ability to detect acute infections during outbreaks. For this purpose, only samples from 2016 were analyzed, when DENV-1 accounted for nearly all circulating strains in the region, to minimize serotype-related differences in antigen detection.

### Dengue Diagnostic Tests and Serotyping

Viremia was assessed by detecting dengue virus-specific NS1 antigen (NS1 Ag) using RDT (SD BIOLINE Dengue DUO kit (NS1 and IgM/IgG), Abbott, IL, USA) and /or viral RNA (DENV RNA) using the RealStar Dengue RT-PCR Kit 3.0 (Altona Diagnostics, Hamburg, Germany). Specific antibodies to DENV (IgM and IgG) were determined by ELISA according to the manufacturer’s instructions (Dia.Pro Diagnostic Bioprobes s.r.l., Milan, Italy). Both assays report sensitivity and specificity above 98%. DENV serotypes were determined using the CDC DENV 1–4 real time PCR assay modified by Santiago et al. [17].

### Classification of the Type of Infection

Given that all serum samples were viremic and collected within the first week after symptom onset, the presence of anti-dengue IgG antibodies was used to classify infection as secondary infections. Among the subset of 107 non-viremic samples, used only for the diagnostic performance evaluation, IgM/IgG ratios and IgG titers were calculated to distinguish primary from secondary infections. Classification of primary and secondary dengue infections using IgG/IgM or IgM/IgG ratios has been widely validated in ELISA-based studies. Samples with low IgM/IgG rates (<1.7) and high IgG antibody titers (above median value= 83 arbU/ml, data not shown) were considered as secondary infections, consistent with the differential kinetics of the humoral response [18, 19].

### Routine Laboratory and Biochemical Assays

Routine laboratory analyses were conducted for all participants in the cohort. Platelet and white blood cell count (WBC) were measured using a CELL-DYN Ruby Hematology Analyzer (Abbott Diagnostics). Hepatic enzyme levels (ALT and AST), as well as total protein, albumin, and bilirubin concentrations, were quantified by photometric methods with the ALINITY Ci-Series system (Abbott Diagnostics).

### Statistical Analysis

Continuous variables were compared using Student’s t-test or the Mann-Whitney U test depending on distribution. Categorical variables were analyzed with Chi-square or Fisher’s exact test. Confidence intervals were set at 95% (CI95); p value<0.05 was considered statistically significant. Concordance between NS1 Ag-RDT and RT-PCR was assessed using Cohen’s kappa coefficient (κ). Agreement was classified as moderate (κ=0.41-0.60), high (κ=0.61-0.80 and almost perfect agreement (κ>0.80). Kappa coefficients were compared using approximate Z-tests based on their standard errors. Statistical analyses were performed using GraphPad Prism 11.0.1.90 software.

### Ethical Aspects

Protocols and procedures were approved by the Ethical Committee of the Academia Nacional de Medicina of Buenos Aires (CEIANM) (TI 12443/16/X; 88/26/CEIANM) and the Ethical Committee of the “Investigación Provincial”, Misiones (CEIP). Written informed consent was obtained from all participants for the use of samples and clinical data in research. Data were stored in a secure, coded database accessible only to study investigators, with personal identifiers replaced by alphanumeric codes to ensure confidentiality.

## RESULTS

### Characteristics of the Study Population

Viremic serum samples from 869 patients were included in this study, comprising 512 collected during 2016 outbreak, 148 from 2019, and 209 from 2024. Sex distribution was similar across the three outbreaks with 288 females (56.25%) in 2016, 79 (53.37%) in 2019, and 116 (55.50%) in 2024 (p=0.1000). Age in years, expressed as median and the interquartile ranges, were 43 (26-67) in 2016, 43 (31-59) in 2019 and 42 (26-58) in 2024 (p=0.8000).

### DENV Serotyping Results

A total of 333 samples were serotyped, including 82 from the 2016 outbreak, 84 from 2019, and 167 from 2024. In 2016, cases were almost exclusively due to DENV-1 [78 (95.12%)], with a small proportion of DENV-4 [4 (4.88%)]. During the 2019, both serotypes circulated again, with DENV-1 remaining predominant [51 (60.71%) were DENV-1 and 33 (39.29%), DENV-4]. In 2024, DENV-2 became predominant [163 (97.60%)], while only a few cases were attributed to DENV-1 [4 (2.40%)].

### Secondary Infection Rates in Viremic Population across Outbreaks

Secondary infections were determined when virus was present concurrently with anti-DENV IgG antibodies. Among the 869 patients analyzed, 535 (61.57%) were classified as having primary infection, while 334 (38.43%) showed evidence of secondary infection. The proportion of secondary infections increased progressively across outbreaks, rising from 31.05% (159 of 512) in 2016 to 43.24% in 2019 (64 of 148), to 53.87% (111 of 214) in 2024 (p < 0.0001).

### Comparison of Clinical Profiles in Primary and Secondary Infections overall and Stratified by Outbreak

Across the three outbreaks, comparison of clinical parameters between primary and secondary infections revealed that secondary infections had significantly higher hematocrit and hemoglobin levels, lower platelet counts (155,000 vs. 140,000 cells/mm³; *p* < 0.0001), and elevated liver enzymes, with higher AST (p < 0.0001) and ALT (p = 0.0003) (Table 1).

**Table 1:**
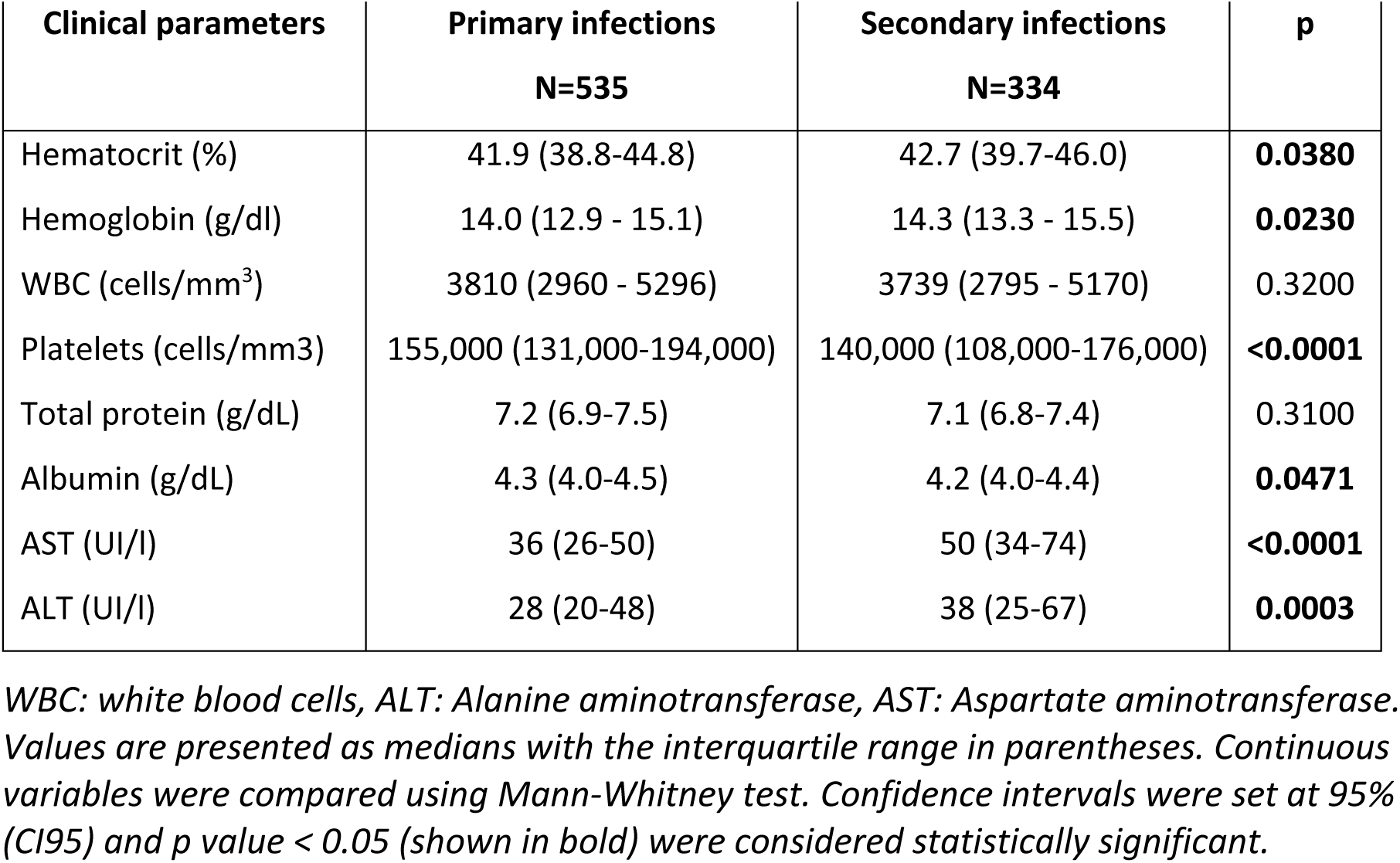
Clinical parameters in primary versus secondary infections in the entire population (N=869)

| Clinical parameters | Primary infections<br>N=535 | Secondary infections<br>N=334 | p |
| --- | --- | --- | --- |
| Hematocrit (%) | 41.9 (38.8-44.8) | 42.7 (39.7-46.0) | **0.0380** |
| Hemoglobin (g/dl) | 14.0 (12.9 - 15.1) | 14.3 (13.3 - 15.5) | **0.0230** |
| WBC (cells/mm<sup>3</sup>) | 3810 (2960 - 5296) | 3739 (2795 - 5170) | 0.3200 |
| Platelets (cells/mm<sup>3</sup>) | 155,000 (131,000-194,000) | 140,000 (108,000-176,000) | **<0.0001** |
| Total protein (g/dL) | 7.2 (6.9-7.5) | 7.1 (6.8-7.4) | 0.3100 |
| Albumin (g/dL) | 4.3 (4.0-4.5) | 4.2 (4.0-4.4) | **0.0471** |
| AST (UI/l) | 36 (26-50) | 50 (34-74) | **<0.0001** |
| ALT (UI/l) | 28 (20-48) | 38 (25-67) | **0.0003** |

WBC: white blood cells, ALT: Alanine aminotransferase, AST: Aspartate aminotransferase. Values are presented as medians with the interquartile range in parentheses. Continuous variables were compared using Mann-Whitney test. Confidence intervals were set at 95% (CI95) and p value < 0.05 (shown in bold) were considered statistically significant.

When we compared clinical parameters between primary and secondary DENV infections within each outbreak, secondary infections showed consistently higher levels of hepatic enzymes (AST and ALT) in all three outbreaks. In 2016, secondary infections were also associated with significantly lower platelet counts (p= 0.0015) (Supplementary Table). In contrast, during 2019, no additional markers of severity were identified when comparing primary and secondary infections (Supplementary Table). The most pronounced differences were observed in 2024, where secondary infections exhibited a broader profile of severity-related parameters, including significant changes in hematocrit, hemoglobin concentration, WBC, platelet counts, and elevated hepatic enzymes, AST and ALT (Table 2). During this outbreak, 88.3% of the individuals with secondary infections exhibited WBC counts below 5000 and nearly 80% showed elevated AST levels.

**Table 2:** Primary versus secondary in 2024 outbreak.

| Clinical parameters | Primary infections<br>N=98 | Secondary infections<br>N=111 | p |
| --- | --- | --- | --- |
| Hematocrit (%) | 42.1 (38.8-44.9) | 43.8 (41.1-46.2) | **0.0124** |
| Hemoglobin (g/dl) | 14.1 (13.2-15) | 14.8 (13.8-15.7) | **0.0077** |
| WBC (cells/mm<sup>3</sup>) | 3350 (2610-4430) | 2880 (2440-3540) | **0.0213** |
| Platelets (cells/mm<sup>3</sup>) | 170,000 (133,500-209,500) | 140,000 (119,000-176,000) | **0.0029** |
| Total protein (g/dL) | 7.4 (7.0-7.7) | 7.4 (7.1-7.8) | 0.7000 |
| Albumin (g/dL) | 4.5 (4.3-4.7) | 4.4 (4.2-4.8) | 0.6000 |
| AST (UI/l) | 38 (29-50) | 50 (40-74) | **0.0007** |
| ALT (UI/l) | 27 (19-38) | 37 (23-59) | **0.0126** |

WBC: white blood cells, ALT: Alanine aminotransferase, AST: Aspartate aminotransferase. Each value is presented as median with the interquartile range in parentheses. Continuous variables were compared using Mann-Whitney test. Confidence intervals were set at 95% (CI95) and a p value < 0.05 (shown in bold) were considered statistically significant.

To explore the clinical relevance of laboratory-warning signs in severe dengue, we considered four laboratory-warning signs: WBC count < 5,000/μL, platelet count < 100,000/μL, and ALT or AST levels above the upper reference limit (40.0 IU/mL). We evaluated the proportion of patients presenting at least one abnormal parameter during each outbreak. During 2024, a significantly higher proportion of individuals with secondary infections exhibited at least one warning sign (93.6%) compared to those observed in 2016 (85.2%) and 2019 (79.2%), indicating a more pronounced severity profile in the most recent epidemic period (p = 0.0300).

### Impact of Secondary Infections in Diagnosis

A total of 318 paired samples were analyzed to assess the concordance between NS1 Ag detection and PCR DENV-RNA testing and the ability of the methodologies to detect acute infections during outbreaks. DENV was detected by any of the 2 methodologies in 211 of 318 samples (viremic cases were 66.4% overall). A substantial number of samples were detected by a single method only, 56 of 211; 26.5%. Considering RT-PCR as the reference standard, the NS1 RDT showed a sensitivity of 85.6% and a specificity of 78.1%. The positive predictive value was 83.8% and the negative predictive value, 80.5%, resulting in overall accuracy of 82.4%. We observed that secondary infections were associated with a twofold higher risk of discordant results between NS1 Ag and PCR compared with non-secondary infections (including primary infections and non-viremic samples) (RR 2.01; 95% CI 1.23–3.30; p= 0.0069) [Table 3, A)]. Cohen’s kappa coefficient was 0.640, consistent with a moderate-to-high level of agreement between the two diagnostic methods. However, agreement between NS1 Ag-RDT and RT-PCR was significantly higher, κ = 0.759, when considering only the group of non-secondary infections (primary plus non-viremic samples) than in secondary infections, κ=0.457. The difference in kappa was statistically significant (K=0.457 vs K=0.759; Δκ=0.302; Z=3.22; p=0.0013), indicating reduced concordance in the context of secondary dengue infection [Table 3, B)].

**Table 3:** *A) Frequency of NS1 Ag and DENV RNA discordant results with and without secondary infections*

|  | Total | Non-secondary infections<br>(primary infections plus<br>non-viremic samples) | % | Secondary infections | % |
| --- | --- | --- | --- | --- | --- |
| Concordant results | 262 | 153 | 58.4 | 109 | 41.6 |
| Discordant results | 56 | 21 | 37.5 | 35 | 62.5 |

| n=318 | RNA (+) | RNA (-) |
| --- | --- | --- |
| NS1 Ag (+) | 155 | 30 |
| NS1 Ag (-) | 26 | 107 |

**K1= 0.640; IC95%= 0.554 – 0.725; SE=0.044**

b- Samples excluding secondary infections (primary infections and non-viremic samples)
| n= 174 | RNA (+) | RNA (-) |
| --- | --- | --- |
| NS1 Ag (+) | 77 | 9 |
| NS1 Ag (-) | 12 | 76 |

**K2= 0.759; IC95%= 0.662 – 0.855; SE=0.049.**

c- Only samples with secondary infections
| n= 144 | RNA (+) | RNA (-) |
| --- | --- | --- |
| NS1 Ag (+) | 78 | 21 |
| NS1 Ag (-) | 14 | 31 |

**K3= 0.457; IC95%= 0.301 – 0.614; SE=0.08**

When the antibody detection platforms of the RDT were compared with ELISA methodologies, the proportion of positive samples identified by the SD BIOLINE IgM and IgG assay was significantly lower than that detected by the ELISA (DIA.PRO) for both markers (p<0.0001), indicating reduced sensitivity of the rapid test. Agreement between the two methods was moderate for IgM and poor for IgG (for IgM: κ=0.495 SE=0.054, IC95%=0.389-0.602; and for IgG: κ=0.183, SE=0.053, IC95%=0.078-0.287). Because of this low overall performance, analyses by primary versus secondary infection were not considered informative.

## DISCUSSION

This study shows a progressive increase in secondary DENV infections in Posadas, Misiones, across consecutive outbreaks, accompanied by marked shifts in the dominant circulating serotype. Our results suggest that the accumulation of previously exposed individuals, along with the introduction of a new predominant serotype, appeared to contribute to a more severe clinical presentation and reduced diagnostic reliability.

In agreement with national reports [3, 4], the 2016 outbreak was almost exclusively caused by DENV-1, with only minimal circulation of DENV-4. By 2019, although DENV-1 remained predominant, DENV-4 accounted for nearly 40% of cases, indicating greater serotype diversification [5, 6]. However, routine surveillance in Argentina relies on serotyping a limited number of samples [3–7]. In contrast, our study includes a substantially larger number of serotyped cases, providing a more robust and region-specific characterization of viral circulation. In 2024, the epidemiological scenario changed markedly with the introduction of DENV-2, which rapidly became the dominant serotype among symptomatic infections, representing almost 98% of cases [7, 8]

Such serotype shifts are recognized as important drivers of epidemic re-emergence and disease severity [20, 21]. Longitudinal cohort studies have shown that most primary dengue infections are inapparent, allowing the silent accumulation of immunity within endemic populations [22]. Repeated outbreaks therefore increase the proportion of individuals with pre-existing immunity, creating favorable conditions for secondary infections when a new serotype is introduced. Supporting this concept, a recent meta-analysis showed that prior dengue infection significantly increased the risk of severe dengue during heterologous secondary infections, despite reducing the likelihood of virologically confirmed infection [23]. Previous studies have demonstrated that secondary infections, particularly those involving DENV-2 or DENV-3 after prior exposure to a different serotype, are more frequently associated with severe outcomes, whereas DENV-1 and DENV-4 infections tend to be milder [24, 25]. In our cohort, the progressive increase in secondary infections together with the circulation of DENV-2 in 2024 was associated with more frequent laboratory warning signs and a more severe clinical presentation. Approximately half of individuals infected with DENV-2 during the 2024 outbreak had secondary infections, highlighting the potential for increased clinical severity.

In addition, secondary infections alter viral kinetics and antigen availability. Rapid viral clearance driven by pre-existing cross-reactive antibodies and memory immune responses may shorten the diagnostic window for RNA detection while also altering the kinetics and availability of circulating NS1 antigen. Previous studies showed lower median viremia levels in secondary infections compared with primary infections [26–28]. In addition, early immune complexes formation may mask NS1 epitopes targeted by diagnostic assays, reducing test sensitivity despite ongoing infection [27, 29, 30].

In our study, secondary infections were associated with reduced concordance between NS1 Ag detection and DENV RNA by RT-PCR, making acute DENV diagnosis more challenging in this immunological context. As a result, a proportion of acute infections during outbreaks may remain undetected depending on the diagnostic methodology used.

Taken together, our findings suggest that the combined use of RT-PCR and NS1 Ag-RDT, along with anti-DENV IgM determination, may be necessary to improve detection of acute DENV infections, particularly in settings with a high frequency of secondary infections. Relying on a single diagnostic approach may underestimate the true number of cases, whereas the complementary use of both methods improves overall diagnostic sensitivity.

### Limitations

Despite the strengths of our study, several limitations should be acknowledged. First, although we analyzed a large number of samples across multiple outbreaks within the same population in Posadas, sequential samples from individual patients were not available, precluding a detailed evaluation of the temporal kinetics of viremia and NS1 antigen levels. Second, the main analysis of secondary infections was restricted to viremic-phase samples, which may have led to underrepresentation of secondary infections occurring outside this window. Third, we were unable to determine the serotype of the primary infection at the individual level; however, based on national surveillance data showing the predominance of DENV-1 during earlier outbreaks in the region, most secondary infections likely corresponded to individuals previously exposed to DENV-1.

## Conclusion

Consecutive outbreaks in Misiones revealed serotype replacement, DENV-2 emergence, and a rising burden of secondary infections associated with greater severity and reduced NS1/PCR concordance. These findings highlight how population immunity influences both disease expression and diagnostic performance supporting the need for integrated diagnostic approaches and tailored therapeutic strategies in highly exposed populations.

## Author contributions

SLE, FAD and PB: Conceptualized and designed the study, collected, and validated data, performed statistical analysis and data interpretation, organized, and curated the dataset, wrote the initial draft of the manuscript and its revision, created and approved the final version to be submitted. MJP, AK, JML, GMC, NA: performed data analysis, conducted dengue diagnostic assays, collected and validated data, reviewed and approved the final version of the manuscript to be submitted. MNB, MM: performed data analysis, organized, and curated the dataset, reviewed the article critically for important intellectual content, approved the final version of the manuscript to be submitted.

## Data availability statement

The data that support the findings of this study are available from the corresponding author upon reasonable request.

## Ethics statement

The study was conducted in accordance with the Declaration of Helsinki and approved by the Institutional Ethics Committee of the Academia Nacional de Medicina, Buenos Aires (TI 12443/16/X; TI N°13157/19/X; 88/26/CEIANM) and the Ethical Committee of the Investigación Provincial, Misiones (CEIP). Informed consent was obtained from all the individuals.

## Acknowledgements

We would like to express our sincere gratitude to all the patients who participated in this study. Their contribution is invaluable to advancing our understanding of dengue virus infections and improving public health outcomes. MJP, FJ, MM, SLE, FAD and PB are members of the National Research Council (CONICET).

## Declaration of generative AI and AI-assisted technologies in the writing process

Artificial intelligence (AI)-based tools (ChatGPT, OpenAI) were used to assist in scientific writing and Kappa exploratory statistical comparisons. All statistical approaches, interpretations, and conclusions were independently reviewed and validated by the authors.

---

**Supplementary Table:** Primary versus secondary in 2016 and 2019 outbreaks.

| Clinical parameters<br>2016 | Primary infections<br>N=353 | Secondary infections<br>N=159 | p |
| --- | --- | --- | --- |
| Hematocrit (%) | 41.0 (38.1-44.7) | 41.8 (38.6-45.5) | 0.2675 |
| Hemoglobin (g/dl) | 13.7 (12.7-15.1) | 14.0 (13.1-15.5) | 0.1772 |
| WBC (cells/mm<sup>3</sup>) | 4290 (3033-5828) | 4500 (3648-5373) | 0.4148 |
| Platelets (cells/mm<sup>3</sup>) | 154,000 (126,500-190,250) | 127,500 (97,875-169,250) | **0.0015** |
| Total protein (g/dL) | 7.1 (6.1-7.4) | 6.9 (6.7-7.3) | 0.1549 |
| Albumin (g/dL) | 4.19 (4.0-4.4) | 4.1(3.8-4.2) | **0.0245** |
| AST (UI/l) | 34 (26-50) | 45 (33-76) | **0.0030** |
| ALT (UI/l) | 29 (22-52) | 40 (27-64) | **0.0176** |
| Clinical parameters<br>2019 | Primary infections<br>N=84 | Secondary infections<br>N=64 | p |
| Hematocrit (%) | 41.9 (39.4-44.7) | 42.1 (39.7-44.8) | 0.5862 |
| Hemoglobin (g/dl) | 14.1 (13.1-15.1) | 14.1 (13.3-15.0) | 0.9900 |
| WBC (cells/mm<sup>3</sup>) | 3929 (3266-5580) | 3760 (2901-5640) | 0.6154 |
| Platelets (cells/mm<sup>3</sup>) | 156,400 (142,500-206,100) | 150,000 (112,450-192,500) | 0.0667 |
| Total protein (g/dL) | 7.1 (6.9-7.6) | 6.9 (6.6 -7.2) | **0.0464** |
| Albumin (g/dL) | 4.3 (4.0-4.5) | 4. 2 (3.9-4.4) | 0.1266 |
| AST (UI/l) | 30 (22-47) | 51 (26-81) | **0.0175** |
| ALT (UI/l) | 30 (18-51) | 43 (27-90) | **0.0094** |

WBC: white blood cells, ALT: Alanine aminotransferase, AST: Aspartate aminotransferase. Each value is presented as median with the interquartile range in parentheses. Continuous variables were compared using Mann-Whitney test. Confidence intervals were set at 95% (CI95) and a p value < 0.05 (shown in bold) were considered statistically significant.

## REFERENCES

1. World Health Organization. Dengue and severe dengue. WHO Fact Sheets. 2025. Available from: https://www.who.int/news-room/fact-sheets/detail/dengue-and-severe-dengue

2. PAHO: Americas report record dengue and Oropouche cases. BMJ. 2024;387:q2808. doi:10.1136/bmj.q2808.

3. Ministerio de Salud de la Nación Argentina. Boletín Integrado de Vigilancia. N°340, SE 51; 2016. Available from: https://www.argentina.gob.ar/sites/default/files/boletin_integrado_vigilancia_n340-se51.pdf

4. Ministerio de Salud de la Nación Argentina. Boletín Integrado de Vigilancia. N°391, SE 51; 2017. Available from: https://www.argentina.gob.ar/sites/default/files/biv_n391-se51.pdf

5. Flichman DM, Pereson MJ, Baré P, Espindola SL, Carballo GM, Albrecht A, Agote F, Alter A, Bartoli S, Blanco S, Blejer J, Borda M, Bouzon N, Carrizo LH, Etcheverry L, Fernandez R, Reyes MIF, Gallego S, Hahn R, Luna SG, Marranzino G, Romanazzi JS, Rossi A, Troffe A, Lin CC, Martínez AP, García G, Di Lello FA. Epidemiology of dengue in Argentina: antibodies seroprevalence in blood donors and circulating serotypes. J Clin Virol. 2022;147:105078. doi:10.1016/j.jcv.2022.105078.

6. Ministerio de Salud de la Nación Argentina. Boletín Integrado de Vigilancia. N°495, SE 19; 2020. Available from: https://bancos.salud.gob.ar/recurso/boletin-integrado-de-vigilancia-n495-se19-2020

7. Ministerio de Salud de la Nación Argentina. Boletín Epidemiológico Nacional. N°711, SE 26; 2024. Available from: https://www.argentina.gob.ar/sites/default/files/2024/04/ben-711-se26.pdf

8. Pan American Health Organization. Informe de situación No. 23. Situación epidemiológica del dengue en las Américas. Published June 27, 2024. Available from: https://www.paho.org/sites/default/files/2024-06/2024-junio-18-phe-actualizacion-dengue-es-final2.pdf

9. Katzelnick LC, Gresh L, Halloran ME, Mercado JC, Kuan G, Gordon A, Balmaseda A, Harris E. Antibody-dependent enhancement of severe dengue disease in humans. Science. 2017;358(6365):929–932. doi:10.1126/science.aan6836.

10. Teo A, Tan HD, Loy T, Chia PY, Chua CLL. Understanding antibody-dependent enhancement in dengue: are afucosylated IgG1s a concern? PLoS Pathog. 2023;19(3):e1011223. doi:10.1371/journal.ppat.1011223.

11. Shih HI, Wang YC, Wang YP, Chi CY, Chien YW. Risk of severe dengue during secondary infection: a population-based cohort study in Taiwan. J Microbiol Immunol Infect. 2024;57(5):730–738. doi:10.1016/j.jmii.2024.07.004.

12. Ghosh A, Mondal S, Sadhukhan S, Sadhukhan PC. Dengue virus and the host immune system: a battle of immune modulation, response and evasion. Pathogens. 2025;14(11):1132. doi:10.3390/pathogens14111132.

13. Liu LT, Huang SY, Lin CH, Chen CH, Tsai CY, Lin PC, Tsai JJ. The epidemiology and identification of risk factors associated with severe dengue during the 2023 dengue outbreak in Kaohsiung City, Taiwan. Travel Med Infect Dis. 2025;65:102852. doi:10.1016/j.tmaid.2025.102852.

14. Blacksell SD. Commercial dengue rapid diagnostic tests for point-of-care application: recent evaluations and future needs. J Biomed Biotechnol. 2012;2012:151967. doi:10.1155/2012/151967.

15. Centers for Disease Control and Prevention. Dengue: NS1 antigen tests for dengue virus. Available from: https://www.cdc.gov/dengue/hcp/diagnosis-testing/dengue-virus-antigen-detection

16. Centers for Disease Control and Prevention. Clinical testing guidance for dengue. Available from: https://www.cdc.gov/dengue/hcp/diagnosis-testing/

17. Santiago GA, Vergne E, Quiles Y, Cosme J, Vazquez J, Medina JF, Medina F, Colón C, Margolis H, Muñoz-Jordán JL. Analytical and clinical performance of the CDC real-time RT-PCR assay for detection and typing of dengue virus. PLoS Negl Trop Dis. 2013;7(7):e2311. doi:10.1371/journal.pntd.0002311.

18. Agarwal A, Jain RK, Chaurasia D, Biswas D. Determining the optimum cut-off IgM/IgG ratio for predicting secondary dengue infections: an observational hospital-based study from Central India. Indian J Med Microbiol. 2022;40(4):492–495. doi:10.1016/j.ijmmb.2022.08.015.

19. Kalra C, Mittal G, Gupta P, Agarwal RK, Ahmad S. Role of IgM/IgG ratio in distinguishing primary and secondary dengue viral infections: a cross-sectional study. Cureus. 2024;16(8):e66714. doi:10.7759/cureus.66714.

20. Ramos-Castañeda J, Barreto Dos Santos F, Martínez-Vega R, Galvão de Araujo JM, Joint G, Sarti E. Dengue in Latin America: Systematic Review of Molecular Epidemiological Trends. PLoS Negl Trop Dis. 2017 Jan 9;11(1):e0005224. doi: 10.1371/journal.pntd.0005224. PMID: 28068335; PMCID: PMC5221820.

21. Sacchetto L, Bernardi V, Brancini ML, Marques BC, Negri A, Vasilakis N, Estofolete CF, Nogueira ML. Early insights of dengue virus serotype 3 (DENV-3) re-emergence in São Paulo, Brazil. J Clin Virol. 2025 Feb;176:105763. doi: 10.1016/j.jcv.2025.105763. Epub 2025 Jan 15. PMID: 39848015.

22. Bos S, Zambrana JV, Duarte E, Graber AL, Huffaker J, Montenegro C, Premkumar L, Gordon A, Kuan G, Balmaseda A, Harris E. Serotype-specific epidemiological patterns of inapparent versus symptomatic primary dengue virus infections: a 17-year cohort study in Nicaragua. Lancet Infect Dis. 2025;25(3):346–356. doi:10.1016/S1473-3099(24)00566-8.

23. Macchia A, Figar S, Biscayart C, González Bernaldo de Quirós F. Impact of prior dengue infection on severity and outcomes: meta-analysis of placebo-controlled trials. Rev Panam Salud Publica. 2024 Dec 4;48:e129. doi: 10.26633/RPSP.2024.129. PMID: 39633829; PMCID: PMC11616458.

24. Vicente CR, Herbinger KH, Fröschl G, Malta Romano C, de Souza Areias Cabidelle A, Cerutti Junior C. Serotype influences on dengue severity: a cross-sectional study on 485 confirmed dengue cases in Vitória, Brazil. BMC Infect Dis. 2016 Jul 8;16:320. doi: 10.1186/s12879-016-1668-y. PMID: 27393011; PMCID: PMC4938938.

25. Soo KM, Khalid B, Ching SM, Chee HY. Meta-Analysis of Dengue Severity during Infection by Different Dengue Virus Serotypes in Primary and Secondary Infections. PLoS One. 2016 May 23;11(5):e0154760. doi: 10.1371/journal.pone.0154760. PMID: 27213782; PMCID: PMC4877104.

26. Duyen HT, Ngoc TV, Ha DT, Hang VT, Kieu NT, Young PR, Farrar JJ, Simmons CP, Wolbers M, Wills BA. Kinetics of plasma viremia and soluble nonstructural protein 1 concentrations in dengue: differential effects according to serotype and immune status. J Infect Dis. 2011;203(9):1292–1300. doi:10.1093/infdis/jir014.

27. Tricou V, Vu HT, Quynh NV, Nguyen CV, Tran HT, Farrar J, Wills B, Simmons CP. Comparison of two dengue NS1 rapid tests for sensitivity, specificity and relationship to viraemia and antibody responses. BMC Infect Dis. 2010;10:142. doi:10.1186/1471-2334-10-142.

28. Bhatt P, Jayaram A, Varma M, Mukhopadhyay C. Kinetics of dengue viremia and its association with disease severity: an ambispective study. VirusDisease. 2024;35(2):250–259. doi:10.1007/s13337-024-00872-z.

29. Muller DA, Choo JJY, McElnea C, Duyen HTL, Wills B, Young PR. Kinetics of NS1 and anti-NS1 IgG following dengue infection reveals likely early formation of immune complexes in secondary infected patients. Sci Rep. 2025;15(1):6684. doi:10.1038/s41598-025-91099-5.

30. Hermann LL, et al. Evaluation of a dengue NS1 antigen detection assay sensitivity and specificity for the diagnosis of acute dengue virus infection. PLoS Negl Trop Dis. 2014;8(10):e3193. doi:10.1371/journal.pntd.0003193.

